# Poor antibody response to BioNTech/Pfizer COVID-19 vaccination in SARS-CoV-2 naïve residents of nursing homes

**DOI:** 10.1101/2021.06.08.21258366

**Authors:** Pieter Pannus, Kristof Y Neven, Stéphane De Craeye, Leo Heyndrickx, Sara Vande Kerckhove, Daphnée Georges, Johan Michiels, Antoine Francotte, Marc Van Den Bulcke, Maan Zrein, Steven Van Gucht, Marie-Noëlle Schmickler, Mathieu Verbrugghe, André Matagne, Isabelle Thomas, Katelijne Dierick, Joshua A. Weiner, Margaret E. Ackerman, Stanislas Goriely, Maria E Goossens, Kevin K. Ariën, Isabelle Desombere, Arnaud Marchant

## Abstract

**Background:** Residents of nursing homes (NH) are at high risk of COVID-19 related morbidity and death and may respond poorly to vaccination because of old age and frequent comorbidities.

**Methods:** Forty residents and forty staff members either naïve or previously infected with SARS-CoV-2 were recruited in two NH in Belgium before immunization with two doses of 30µg BNT162b2 mRNA vaccine at day 0 and day 21. Binding antibodies (Ab) to SARS-CoV-2 receptor binding domain (RBD), spike domains S1 and S2, RBD Ab avidity, and neutralizing Ab against SARS-CoV-2 wild type and B.1.351 variant were assessed at days 0, 21, 28, and 49.

**Results:** SARS-CoV-2 naïve residents had lower Ab responses to BNT162b2 mRNA vaccination than naïve staff. These poor responses involved lower levels of IgG to all domains of the vaccine antigen, lower avidity of RBD IgG, and lower levels of Ab neutralizing the vaccine strain. No naïve resident had detectable neutralizing Ab to the B.1.351 variant. High and comparable Ab responses were observed in residents and staff previously infected with SARS-CoV-2. Clustering analysis revealed that poor vaccine responders not only included naïve residents but also naïve staff, emphasizing the heterogeneity of responses to mRNA vaccination in the general population.

**Conclusions:** The poor Ab responses to mRNA vaccination observed in infection naïve residents and in some naïve staff members of NH suggest suboptimal protection against breakthrough infection, especially with variants of concern. Adapted vaccination regimens may be needed to provide optimal protection against COVID-19 to vulnerable populations.

**Summary:** Poor antibody responses to COVID-19 mRNA vaccination were observed in SARS-CoV-2 infection naïve residents and some naïve staff members of nursing homes. This suggests suboptimal protection against breakthrough infection, especially with variants of concern, and the need for adapted vaccination regimens.

## Introduction

Residents of nursing homes (NH) are at a disproportionately high risk of COVID-19 related morbidity and mortality, representing about 5% of all cases while accounting for >30% of all COVID-19 related deaths in the United States [1,2]. Vaccination campaigns around the world have therefore generally prioritized NHs, achieving high coverage rates especially among residents [3,4]. As a result, new cases and deaths have declined steeply in such facilities, outpacing national rates [5–7].

The success of COVID-19 mRNA vaccination in NH is consistent with data from phase 2 studies indicating potent immunogenicity of these vaccines in younger and older adults [8,9]. However, recent observational studies have found lower antibody (Ab) responses to BNT162b2 vaccination in older adults [10–12]. In addition, chronic comorbidities such as type 2 diabetes and cardiovascular disease were associated with lower vaccine responses [11,13]. These data raise the concern that NH residents, who are old, frail, and often have comorbidities, might respond more poorly to COVID-19 vaccination. Supporting this concern, a retrospective observational cohort study from Denmark found lower vaccine effectiveness in NH residents (64%) as compared to healthcare workers (90%) one week after the second dose of BNT162b2 mRNA vaccination [14].

Decreased efficacy of COVID-19 vaccination in NH residents may be particularly problematic in the face of emerging SARS-CoV-2 variants that are less susceptible to vaccine-induced neutralizing Ab [15–17]. Breakthrough infections with SARS-CoV-2 variants following complete mRNA vaccination have been reported in healthy adults and more recently, severe COVID-19 and death have been reported following an outbreak of the SARS-CoV-2 R.1 variant in a NH in Kentucky [18,19]. The concern of severe breakthrough infection with SARS-CoV-2 variants may be lower in NH residents who have survived natural infection. Indeed, COVID-19 mRNA vaccination induces higher Ab responses in previously infected adults as compared to infection naïve adults and boosts neutralizing Ab cross-reacting with variants of concern [20–25]. The level of cross-reactive immunity induced by mRNA vaccination in naïve and previously infected NH residents is currently unclear.

Taken together, available data raise concern regarding immunity induced by current COVID-19 mRNA vaccine regimens in infection naïve and frail NH residents, especially in the current context of emerging SARS-CoV-2 variants. We therefore established a longitudinal cohort of SARS-CoV-2 naïve or previously infected NH residents and staff who received two doses of the BNT162b2 mRNA vaccine and assessed the magnitude and quality of Ab responses to SARS-CoV-2 Wuhan (wild type, WT) and B.1.351, first identified in South Africa, as a prototype variant of concern.

## Material and methods

### Study design and approvals

This study is nested in a prospective cohort study named PICOV (Prior Infection with SARS-CoV-2) [26]. The objective of this nested study was to measure the immune response to SARS-CoV-2 mRNA vaccination in naïve and previously infected residents and members of staff. The study was approved by the Ethics Committee of the Hôpital Erasme, Brussels, Belgium (reference B4062020000134), the Federal Agency for Medicines and Health Products (2021-000401-24), and is registered on ClinicalTrials.gov (NCT04527614).

### Recruitment and clinical sample collection

SARS-CoV-2 infection-naïve and previously infected residents and staff from two Belgian NHs were recruited. Those with a documented positive RT-qPCR or serological test result at baseline were considered to be previously infected with SARS-CoV-2. Main exclusion criteria for NH residents included a previous diagnosis of dementia, a mini-mental state examination (MMSE) score ≤18/30, and life expectancy <6 months. As described previously, scores from the Clinical Frailty Scale (CFS) and Quality of Life (QoL) were determined for residents at baseline [26].

All subjects were immunized with two doses of 30μg BNT162b2 mRNA from BioNTech/Pfizer (Comirnaty®), 21 days apart. Blood samples were collected on the day of the primary dose (baseline or day 0), the day of the boost (day 21) as well as one and four weeks after the boost (respectively day 28 and day 49). Serum was separated by blood centrifugation at 1000g for 10 minutes and stored at -20°C for downstream Ab analyses.

### SARS-CoV-2-Specific Binding Antibodies

Levels of serum Ab were assessed using a multiplexed immunoassay (Multi-SARS-CoV-2 Immunoassay), developed in collaboration with InfYnity Biomarkers (Lyon, France). In this microarray, SARS-CoV-2 antigens, selected for their individual performance, were printed in 96-well polystyrene microplates using a sciFLEXARRAYER printing system (Scienion, Germany). Individual SARS-CoV-2 antigens, including Spike 1 domain (S1, encompassing AA16-685 of S), Spike 2 domain (S2, encompassing AA686-1213 of S), and Receptor Binding Domain (RBD), were printed in duplicate (GenBank YP009724390.1). Serial dilutions of test samples as well as positive and negative control sera were incubated in microarray plates for 1h at room temperature (RT) and washed with phosphate-buffered saline with 0.05% Tween 20 (PBST). Next, plates were incubated (1h, RT) with horseradish peroxidase-conjugated goat anti-human IgG and washed with PBST before adding a precipitating TMB solution for 20min (RT, dark). Then, TMB was removed and plates were dried at 37°C for 10min. Microplates were imaged and analyzed using a microplate reader (SciReader CL, Scienion, Germany). The average pixel intensity for each spot was calculated for each antigen/dilution and reported as net intensity. The dynamic range of each antigen measurement was defined using serial dilutions of positive sera. Only antigen measurements within the dynamic range were considered and were multiplied by the dilution factor. For each serum, quantitative results were eligible if at least 2 dilutions report comparable results (%CV<28%). Results are reported as arbitrary pixel units per milliliter (AU/ml). ROC-analyses using an independent population for validation generated cutoff concentrations of 21.0 AU/ml, 19.5 AU/ml and 19.5 AU/ml for RBD, S1 and S2, respectively (**Supplementary methods**).

### SARS-CoV-2 Neutralizing Antibodies

Serial dilutions of heat-inactivated serum (1/50-1/25600 in EMEM supplemented with 2mM L-glutamine, 100U/ml - 100μg/ml of Penicillin-Streptomycin and 2% fetal bovine serum) were incubated during 1h (37°C, 7% CO_2_) with 3xTCID100 of (i) a wild type (WT) Wuhan strain (2019-nCoV-Italy-INMI1, reference 008V-03893) and (ii) the B.1.351 variant of SARS-CoV-2, in parallel. Sample-virus mixtures and virus/cell controls were added to Vero cells (18.000 cells/well) in a 96-well plate and incubated for five days (37°C, 7% CO_2_). The cytopathic effect caused by viral growth was scored microscopically. The Reed-Muench method was used to calculate the neutralizing Ab titer that reduced the number of infected wells by 50% (NT50), which was used as a proxy for the neutralizing Ab concentration in the sample [27].

### SARS-CoV-2 RBD-Specific antibody avidity

Bio-layer interferometry measurements were performed with an Octet HTX instrument (Fortébio) using AR2G biosensors. Data analyses were performed using FortéBio Data Analysis 9.0 software. Kinetic assays were performed at 25-30°C at a sample plate agitation speed of 1000rpm. Sensors were first activated by immersion in a solution containing 20mM EDC and 10mM s-NHS. Then, 0.05mg/ml of RBD antigen in 10mM sodium acetate pH 6 was loaded for 600sec. After antigen loading, the biosensors were immersed in a solution of 1M ethanolamine pH8.5 to prevent non-specific interactions. Antigen loaded AR2G sensors were first dipped in PBS to establish a baseline time curve, and then immersed for 10min in wells containing purified serum IgG at three different dilutions (3-5-8x). Following IgG association, dissociation was monitored for 600sec in PBS. Negative controls included ligand without IgG and IgG without ligand. Kinetic parameters were determined by global fitting of the association and dissociation phases of the binding curves according to a 1:1 binding model.

### Statistical analyses

Analyses were performed in R (version 4.0.3). Categorical data were presented as frequencies and percentages, continuous data as means (SD). The Kruskall-Wallis test and post-hoc Mann-Whitney U test alongside multiple testing correction with the false discovery rate were used for all time wise group comparisons. The Mann-Whitney test was used to compare WT and B.1.351 variant neutralizing Ab at day 49. Spearman’s rank correlation coefficients (rho, ρ) were determined for associations between WT and B.1.351 variant neutralizing Ab, SARS-CoV-2 binding Ab, and Ab avidity.

A Uniform Manifold Approximation and Projection (UMAP) analysis was performed using the R package “umap” for dimensionality reduction of the following outcomes at day 49: anti-RBD/S1/S2 IgG, anti-RBD IgG avidity, and WT NT50. To achieve normality, avidity was log_10_ and neutralization log_2_ transformed. The optimal number of clusters was tested via the k-means (range 1:10) and visually identified with an “elbow” in a plot of variance versus number of clusters. DBSCAN (“dbscan” package) identified clusters within the UMAP reduced dimensions.

## Results

The study included 40 residents and 40 members of staff who were either naïve or previously infected with SARS-CoV-2 before they received 2 x 30µg BNT162b2 mRNA vaccine at their respective NH. In previously infected subjects, SARS-CoV-2 infection occurred between 269 and 315 days before vaccination. Complete cohort and demographic information is provided in **Table 1**. Although residents with the poorest health status were excluded, most enrolled residents were frail and many suffered multiple co-morbidities requiring medication.

**Table 1.** Demographic Characteristics of the Participants, According to Study Group.

|  | naive staff<br>(N=19) | naive resident<br>(N=20) | infected<br>staff<br>(N=21) | infected<br>resident<br>(N=20) | Total<br>(N=80) | p value |
| --- | --- | --- | --- | --- | --- | --- |
| **Age, years** |  |  |  |  |  | <0.001 |
| Mean (SD) | 47.9 (10.2) | 86.0 (8.3) | 47.6 (11.0) | 84.3 (7.7) | 66.4 (21.0) |  |
| Range | 23.0 - 64.0 | 67.0 - 102.0 | 30.0 - 68.0 | 65.0 - 94.0 | 23.0 - 102.0 |  |
| **Gender** |  |  |  |  |  | 0.66 |
| Female | 14 (73.7%) | 13 (65.0%) | 16 (76.2%) | 12 (60.0%) | 55 (68.8%) |  |
| Male | 5 (26.3%) | 7 (35.0%) | 5 (23.8%) | 8 (40.0%) | 25 (31.2%) |  |
| **Ethnicity** |  |  |  |  |  | 0.13 |
| Caucasian | 17 (89.5%) | 20 (100.0%) | 18 (85.7%) | 20 (100.0%) | 75 (93.8%) |  |
| Other | 2 (10.5%) | 0 (0.0%) | 3 (14.3%) | 0 (0.0%) | 5 (6.2%) |  |
| **BMI, kg/m<sup>2</sup> \*** |  |  |  |  |  | 0.009 |
| Mean (SD) | 27.3 (5.3) | 24.8 (6.0) | 28.1 (5.3) | 22.9 (4.3) | 25.8 (5.6) |  |
| Range | 21.2 - 36.8 | 16.7 - 36.3 | 20.1 - 44.2 | 14.6 - 30.5 | 14.6 - 44.2 |  |
| **Self-reported smoking status** |  |  |  |  |  | 0.48 |
| Ex-smoker | 1 (5.3%) | 1 (5.0%) | 3 (14.3%) | 5 (25.0%) | 10 (12.5%) |  |
| Non-smoker | 16 (84.2%) | 18 (90.0%) | 17 (81.0%) | 14 (70.0%) | 65 (81.2%) |  |
| Current smoker | 2 (10.5%) | 1 (5.0%) | 1 (4.8%) | 1 (5.0%) | 5 (6.2%) |  |
| **Daily exercise** |  |  |  |  |  | 0.005 |
| less than 30 minutes | 3 (15.8%) | 12 (60.0%) | 2 (9.5%) | 10 (50.0%) | 27 (33.8%) |  |
| 30 to 60 minutes | 6 (31.6%) | 6 (30.0%) | 7 (33.3%) | 6 (30.0%) | 25 (31.2%) |  |
| at least 60 minutes | 9 (47.4%) | 2 (10.0%) | 12 (57.1%) | 3 (15.0%) | 26 (32.5%) |  |
| None | 1 (5.3%) | 0 (0.0%) | 0 (0.0%) | 1 (5.0%) | 2 (2.5%) |  |
| **Self-reported health status** |  |  |  |  |  | <0.001 |
| Very good health | 9 (47.4%) | 0 (0.0%) | 4 (19.0%) | 2 (10.0%) | 15 (18.8%) |  |
| Good health | 10 (52.6%) | 12 (60.0%) | 14 (66.7%) | 8 (40.0%) | 44 (55.0%) |  |
| Reasonable health | 0 (0.0%) | 8 (40.0%) | 2 (9.5%) | 9 (45.0%) | 19 (23.8%) |  |
| Bad health | 0 (0.0%) | 0 (0.0%) | 1 (4.8%) | 1 (5.0%) | 2 (2.5%) |  |
| **Quality of Life index** |  |  |  |  |  | <0.001 |
| Mean (SD) | 0.9 (0.1) | 0.6 (0.3) | 0.8 (0.2) | 0.6 (0.2) | 0.7 (0.2) |  |
| Range | 0.7 - 1.0 | 0.1 - 1.0 | 0.5 - 1.0 | 0.2 - 1.0 | 0.1 - 1.0 |  |
| **Medication use** |  |  |  |  |  |  |
| Cardiovascular disease | 1 (5.3%) | 17 (85.0%) | 1 (4.8%) | 19 (95.0%) | 38 (47.5%) | <0.001 |
| Hypertension | 2 (10.5%) | 14 (70.0%) | 3 (14.3%) | 19 (95.0%) | 38 (47.5%) | <0.001 |
| Pain medication | 0 (0.0%) | 15 (75.0%) | 0 (0.0%) | 12 (60.0%) | 27 (33.8%) | <0.001 |
| Diabetes Mellitus | 0 (0.0%) | 4 (20.0%) | 0 (0.0%) | 3 (15.0%) | 7 (8.8%) | 0.05 |
| Anti-psychotic medication | 0 (0.0%) | 10 (50.0%) | 0 (0.0%) | 8 (40.0%) | 18 (22.5%) | <0.001 |
| Anti-depressant medication | 0 (0.0%) | 12 (60.0%) | 0 (0.0%) | 7 (35.0%) | 19 (23.8%) | <0.001 |
| Pulmonary disease | 0 (0.0%) | 3 (15.0%) | 0 (0.0%) | 1 (5.0%) | 4 (5.0%) | 0.10 |
| Allergy | 0 (0.0%) | 2 (10.0%) | 0 (0.0%) | 3 (15.0%) | 5 (6.2%) | 0.12 |
| Neurological disease | 0 (0.0%) | 4 (20.0%) | 0 (0.0%) | 2 (10.0%) | 6 (7.5%) | 0.05 |
| Immunological disease | 0 (0.0%) | 0 (0.0%) | 1 (4.8%) | 0 (0.0%) | 1 (1.2%) | 0.42 |
| **MMSE score<sup>†</sup>** |  |  |  |  |  | 0.98 |
| Mean (SD) | . | 25.6 (3.2) | . | 25.9 (3.3) | 25.8 (3.2) |  |
| Range | . | 19.0 - 30.0 | . | 18.0 - 30.0 | 18.0 - 30.0 |  |
| **Frailty scale<sup>†</sup>** |  |  |  |  |  |  |
| Very fit | . | 0 (0.0%) | . | 1 (5.0%) | 1 (2.5%) | 0.40 |
| Fit | . | 3 (15.0%) | . | 1 (5.0%) | 4 (10.0%) | 0.28 |
| Managing well | . | 5 (25.0%) | . | 7 (35.0%) | 12 (30.0%) | 0.87 |
| Very mild frailty | . | 3 (15.0%) | . | 3 (15.0%) | 6 (15.0%) | 1.00 |
| Mild frailty | . | 6 (30.0%) | . | 4 (20.0%) | 10 (25.0%) | 0.55 |
| Moderate frailty | . | 1 (5.0%) | . | 3 (15.0%) | 4 (10.0%) | 0.47 |
| Severe frailty | . | 2 (10.0%) | . | 1 (5.0%) | 3 (7.5%) | 0.39 |

Data are mean (SD) or n (%). Range denotes the lowest to the highest numerical observation.
\* Data available for 19, 19, 21, 20, and 79 subjects.
† Mini-mental State Examination (MMSE) and Frailty scale was only asked to residents.

Levels of binding Ab to SARS-CoV-2 spike receptor binding domain (RBD), spike subunit domains one (S1) and two (S2) were measured in longitudinal serum samples using a multiplex immunoassay. Detailed numerical data are presented in **Tab.S1**. At baseline, naïve staff and residents had undetectable levels of SARS-CoV-2-specific IgG whereas high levels of Ab were detected in previously infected subjects (**Fig.1a, Fig.S1**). Primary vaccination induced a significant increase in SARS-CoV-2 Ab in naïve as well as previously infected staff and residents, and Ab levels were further boosted following secondary vaccination at day 21 (**Fig.1a**). Vaccine-induced Ab levels to RBD and S1 were about six-fold lower in naïve residents as compared to naïve staff following primary vaccination and two-fold lower after booster vaccination (**Fig.1b**). In comparison to naïve subjects, Ab levels were strongly increased in both residents and staff previously infected with SARS-CoV-2 (**Fig.1b and Fig.S1**). Among previously infected subjects, residents had higher Ab responses to RBD and S1 as compared to staff. Ab responses to S2 were lower than responses to RBD and S1, especially in naïve subjects.

**Figure 1.**
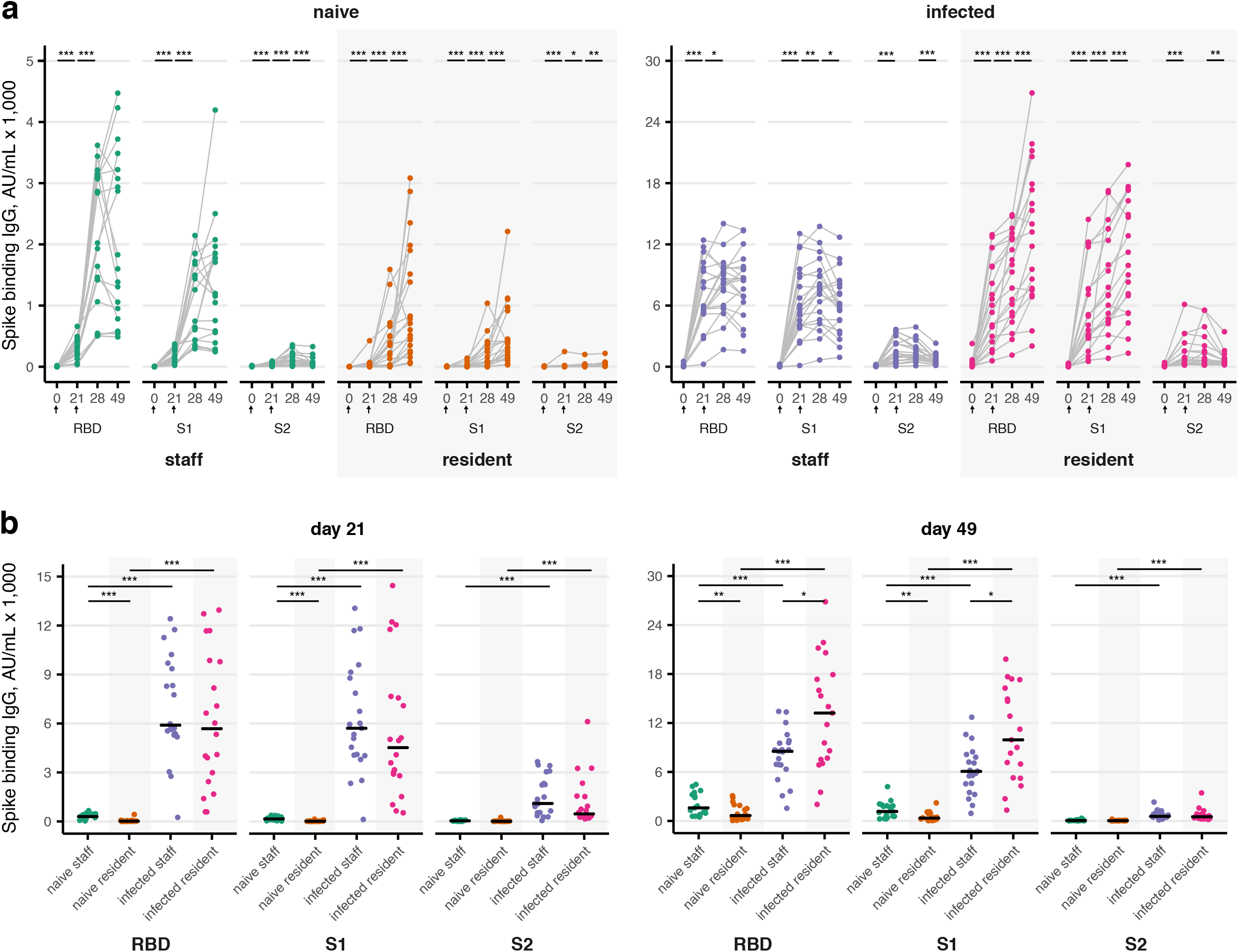
SARS-CoV-2 Specific Binding Antibody Responses to BNT162b2 mRNA Vaccination in Residents and Staff of Nursing Homes. SARS-CoV-2 naïve and previously infected NH residents and staff received two doses of 30µg BNT162b2 vaccine on day 0 and day 21 (arrows). The concentration of spike-specific binding Ab was measured using a multiplex assay before vaccination and at days 21, 28 and 49 after the first dose and is shown as arbitrary pixel units per ml (AU/ml; limit of quantification, 21.0 for RBD, 19.5 for S1 and S2). Each data point represents a serum sample. Statistical significance of differences between time points (**panel A**) and study groups (**panel B**) were determined by the Kruskall-Wallis test by ranks, using the Mann-Whitney U post-hoc test and Benjamini-Hochberg correction for multiple testing (*: p<0.05; **: p<0.01; ***: p<0.001).

The avidity of RBD-specific Ab was measured using bio-layer interferometry. Rapid avidity maturation was observed after primary vaccination in naïve and previously infected staff (**Fig.2a**). High RBD IgG avidity was also observed in previously infected residents at day 21, whereas avidity could only be assessed in few naïve residents who had sufficiently high levels of RBD Ab to be characterized (**Fig.2a**). Following booster vaccination, RBD IgG avidity further increased in naïve staff and residents, but remained stable in previously infected subjects (**Fig.2a**). Four weeks after booster vaccination (day 49), Ab avidity was significantly higher in naïve staff as compared to naïve residents, and was higher in previously infected subjects as compared to naïve subjects (**Fig.2b**).

**Figure 2.**
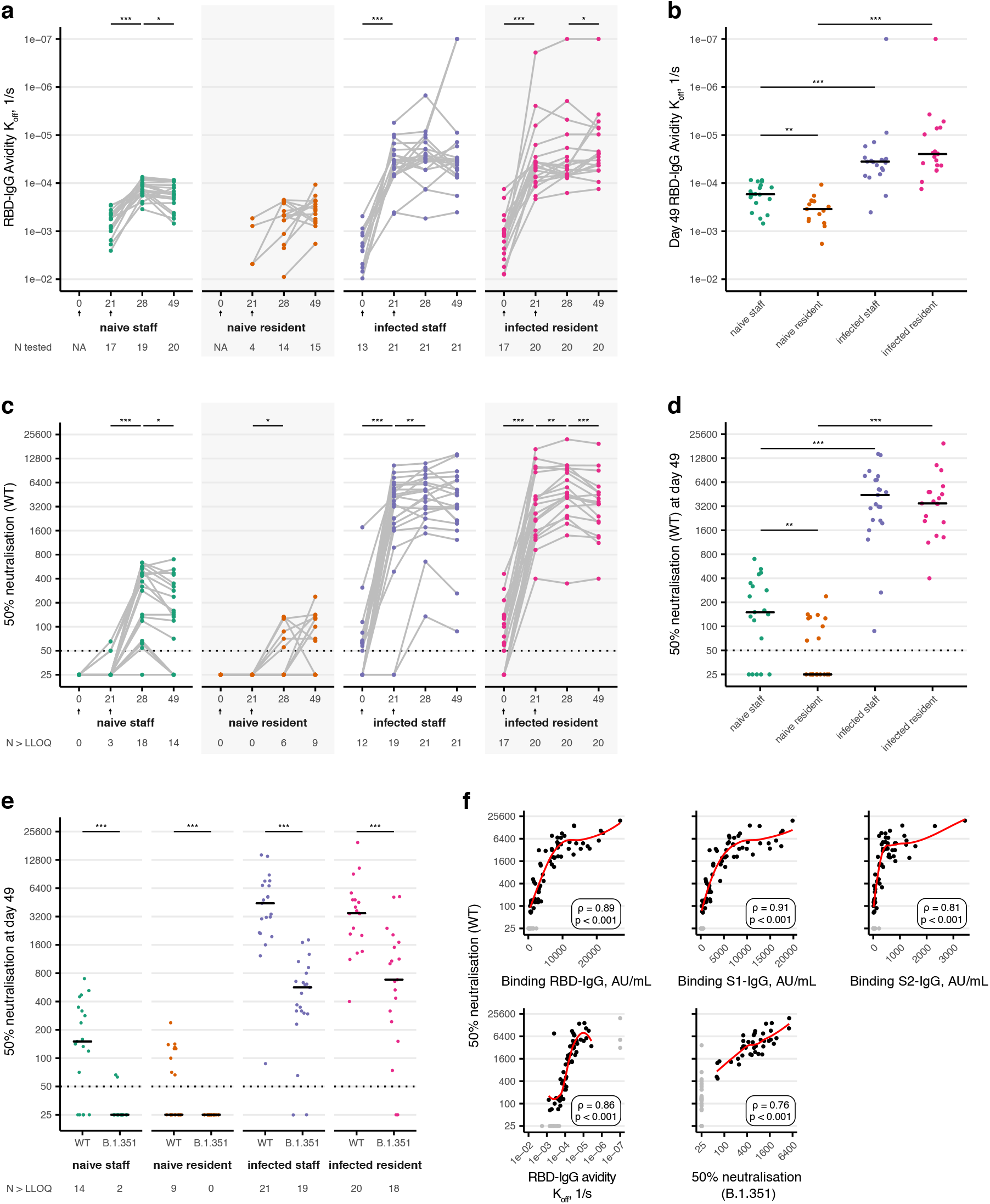
Low RBD IgG Avidity and Neutralizing Antibody Responses in SARS-CoV-2 Naïve Residents. RBD IgG avidity and neutralizing Ab responses to mRNA vaccination were measured at days 0, 21, 28 and 49 in SARS-CoV-2 naïve and previously infected residents and staff. **Panels A and B**. Avidity of RBD-specific IgG (K_off_ in 1/s). ‘N tested’ indicates the number of participants with sufficiently high antibody concentrations for avidity testing (panel A). **Panels C/D/E**. 50% neutralizing Ab titers of SARS-CoV-2 wild type (WT) and B.1351 variant (lower limit of quantification, LLOQ, 1/50). ‘N > LLOQ’ indicates the number of participants with detectable neutralizing Ab (panel C). Black bars indicate median values. Statistical significance of differences between time points and study groups were determined by the Kruskall-Wallis test by ranks, using the Mann-Whitney U post-hoc test and Benjamini-Hochberg correction for multiple testing (*: p<0.05; **: p<0.01; ***: p<0.001). For differences between wild type and the B.1.351 variant the Mann-Whitney test was used. **Panel F**. Spearman’s rank correlation coefficients (rho, ρ) between titers of neutralizing Ab to WT strain and the other Ab response parameters. Data below or above limits of quantification were excluded (gray dots).

The lower levels and avidity of vaccine-induced Ab observed in naïve residents as compared to naïve staff suggested lower neutralizing Ab capacity. To explore this possibility, titers of neutralizing Ab against WT Wuhan strain and B.1.351 variant were measured. Previously infected staff and residents had detectable neutralizing Ab to the Wuhan strain at baseline and these titers further increased by primary and booster vaccinations (**Fig.2c**). Potent neutralizing Ab responses were also induced by vaccination of naïve staff, although the proportion of subjects with detectable responses decreased between day 28 (18/19) and day 49 (14/19). In contrast, only 6/20 naïve residents had detectable neutralizing Ab at day 28 and this proportion increased to 9/20 at day 49 (**Fig.2c**). At day 49, naïve residents had significantly lower neutralizing Ab responses as compared to naïve staff, whereas higher responses were detected in previously infected subjects as compared to naïve subjects (**Fig.2d**). Compared to the wild type strain, neutralizing titers against the B.1.351 variant were reduced five to ten-fold across study groups (**Fig.2e**). In naïve subjects, only 2/19 staff and none of the naïve residents had detectable neutralizing Ab against the B.1.351 variant at day 49, whereas neutralizing Ab were detected in 19/21 previously infected staff and 18/20 previously infected residents.

The consistent differences in Ab responses observed between the four study groups suggested a coordinated response to mRNA vaccination across the measured immunological parameters. Indeed, titers of neutralizing Ab against the wild type strain strongly correlated with RBD, S1 and S2 binding Ab, RBD IgG avidity, and neutralizing Ab to the B.1.351 variant (**Fig.2f**).

To further explore inter-individual variability of this coordinated response, a clustering analysis was performed to reduce the complete dataset to two dimensions and identify groups of subjects who have similar profiles of Ab responses. Five clusters of study participants with distinct Ab levels, avidity, and neutralizing activity at day 49 were identified (**Fig.3a-d**). These clusters were not correlated with age of the study participants (**Fig.3e**). Clusters 4 and 5 exclusively contained previously infected subjects with high Ab responses. Interestingly, cluster 5, including the highest Ab responses, was mostly composed of previously infected residents. In contrast, cluster 1, including the lowest Ab responses, was composed of a mix of naïve residents and naïve staff, indicating that both populations contain low responders to mRNA vaccination. Clusters 2 and 3 included intermediate Ab responses and were composed of a mix of naïve residents, naïve staff and some previously infected staff and residents. The clustering analysis therefore revealed a group of poor Ab responders that not only included naïve residents but also naïve staff.

**Figure 3.**
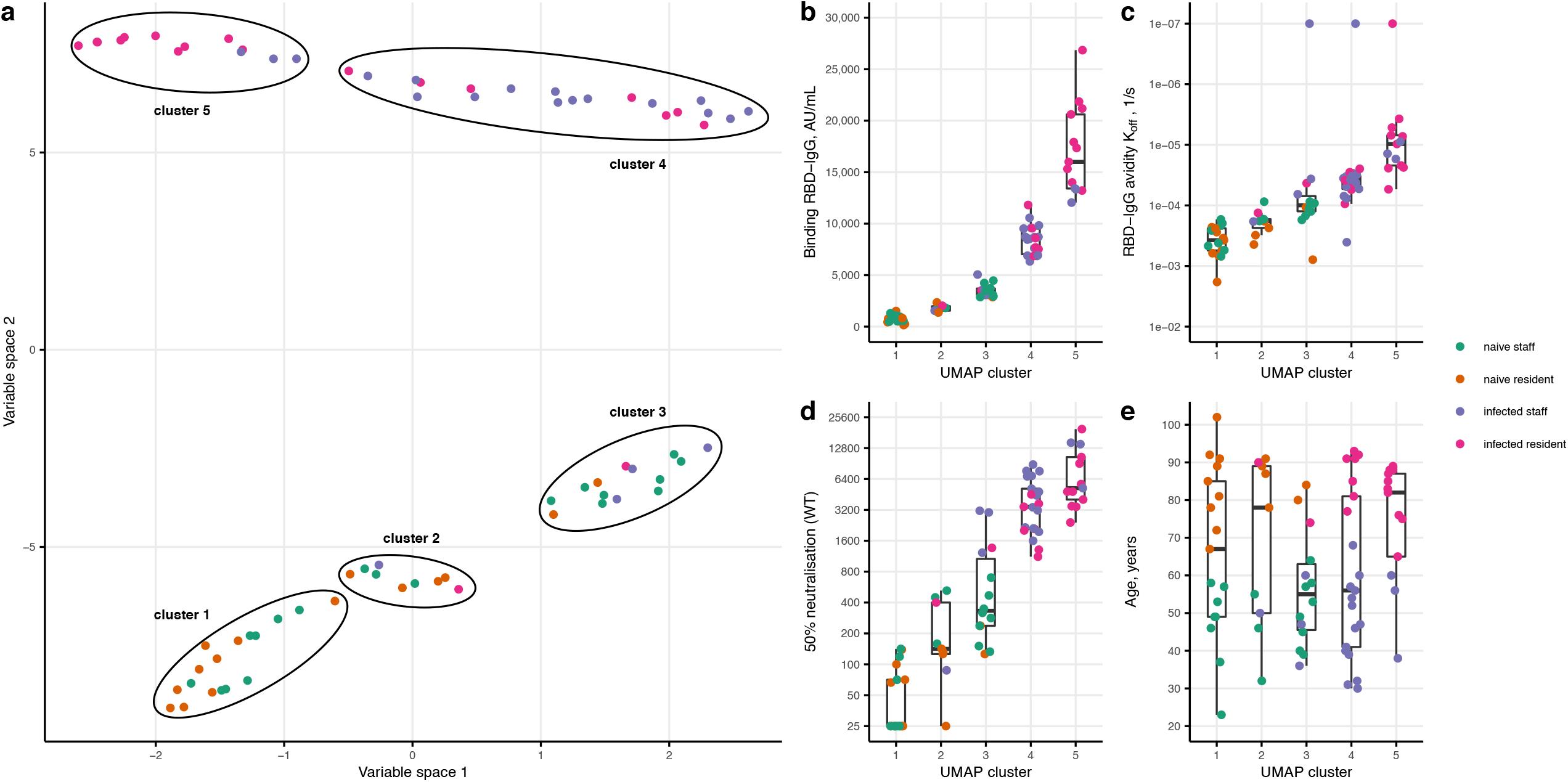
Low Vaccine Responders Include both SARS-CoV-2 Naïve Nursing Home Residents and Staff. **Panel A**. Clustering (UMAP) analysis of all study participants with available RBD/S1/S2 binding IgG Ab concentrations, RBD-IgG avidity and SARS-CoV-2 wild type neutralization at day 49. The position of individual participants in variable space 1 and 2 indicates similarities or differences in Ab responses. DBSCAN was used to identify clusters. **Panels B/C/D**. Clusters 1 to 5 are plotted against the RBD binding IgG, RBD IgG avidity and WT neutralizing titers, respectively. **Panel E**. Age of participants included in clusters of antibody responses. Black bars indicate median values.

## Discussion

Reports on lower Ab responses to COVID-19 mRNA vaccination in older people and in people with chronic comorbidities raise concern about the susceptibility of NH residents to severe breakthrough infections, especially with SARS-CoV-2 variants of concern [10–13]. In this study, NH residents without previous SARS-CoV-2 infection had lower Ab responses to BNT162b2 mRNA vaccination as compared to naïve staff. These defective responses included lower levels of IgG to all domains of the vaccine antigen, lower avidity of RBD IgG and lower levels of neutralizing Ab. Worryingly, none of the naïve residents had detectable neutralizing Ab to the B.1.351 variant.

Although an immune correlate of protection against COVID-19 has not been established yet, levels of virus-specific binding and neutralizing Ab have been shown to correlate with vaccine efficacy in phase 3 studies across different vaccination platforms [28]. In addition, data from pre-clinical studies on non-human primates indicate that mRNA vaccine-induced neutralizing Ab can mediate protection against disease [29–31]. The poor Ab responses observed in our study are therefore likely associated with lower vaccine-induced protection. Providing optimal protection to the vulnerable population of NH residents may require adapted vaccination regimens, such as additional doses of homologous or heterologous vaccines.

Both age and health status differentiate NH residents and staff. In this cohort, Ab responses were not strongly correlated with age, suggesting a more important role of health status, including frailty and comorbidities. This observation is consistent with the robust Ab responses to mRNA vaccination observed in older people with preserved health status and living outside NH [32]. In both residents and staff, previous SARS-CoV-2 infection was a major determinant of Ab responses, with markedly higher Ab levels and quality in previously infected as compared to naïve subjects. NH residents previously infected with SARS-CoV-2 had remarkably high Ab responses to mRNA vaccination and included the highest responders of the cohort. These high vaccine responses likely reflect potent immunological memory potentially induced by more severe infections and selected by survival after COVID-19 [33]. In marked contrast with naïve residents, NH residents previously infected with SARS-CoV-2 may be at particularly low risk of breakthrough infection following mRNA vaccination.

Another important finding of this study is that poor vaccine responders were not limited to naïve residents, but also included healthy naïve staff. This observation emphasizes the heterogeneity of Ab responses to mRNA vaccination in the general population [34–36]. As mRNA vaccination has only recently been implemented in large populations, the immunological basis of this heterogeneity is currently unknown. Systems immunology, involving high dimensional analyses of the immune system, is emerging as a promising approach to identify determinants of vaccine responsiveness and has the potential to guide the development of next-generation mRNA vaccines against COVID-19 and other target pathogens [37,38].

Identifying vulnerable populations who may benefit less from current mRNA vaccination regimens is essential for the control of the COVID-19 pandemic. Adapted mRNA vaccination regimens may be required to protect SARS-CoV-2 naïve residents of NH and younger poor vaccine responders against breakthrough infections, especially with variants of concern.

## Data Availability

All study data is stored on a secured central server of Sciensano.

## Acknowledgements

We thank Caroline Rodeghiero, Fabienne Jurion, Elfriede Heerwegh, Giresse Tima, Elisa Brauns, Muriel Nguyen, Séverine Thomas, Vincent Martens, Valérie Acolty, Véronique Olislaghers, Inès Vu Duc, Betty Willems, Maria Lara Escandell, Sandra Coppens, Ann Ceulemans, Koen Bartholomeeusen and Marylène Vandevenne for their technical and logistical help in the laboratory. We thank Martine Delaere, Kristine Massez, Jody Serré, Nathalie Matia Sangrador and Elodie Glinne for their excellent and dedicated work as study nurses. We thank Dr. Piet Maes (Rega Institute, KU Leuven, Belgium) to kindly provide the B.1.351 SARS-CoV-2 isolate. Finally, we thank all residents and members of staff of the participating nursing homes for their availability, flexibility and dedication to the study. S.G. is Senior research Associate and A.M. is Research Director of the FRS-FNRS, Belgium.

## References

1. Werner RM, Hoffman AK, Coe NB. Long-Term Care Policy after Covid-19 - Solving the Nursing Home Crisis. N Engl J Med 2020; 383:903–905.

2. Apr 23 P, 2021. State COVID-19 Data and Policy Actions. KFF. 2021; Available at: https://www.kff.org/coronavirus-covid-19/issue-brief/state-covid-19-data-and-policy-actions/. Accessed 27 April 2021.

3. Dooling K. The Advisory Committee on Immunization Practices’ Interim Recommendation for Allocating Initial Supplies of COVID-19 Vaccine — United States, 2020. MMWR Morb Mortal Wkly Rep 2020; 69. Available at: https://www.cdc.gov/mmwr/volumes/69/wr/mm6949e1.htm. Accessed 27 April 2021.

4. Gharpure R. Early COVID-19 First-Dose Vaccination Coverage Among Residents and Staff Members of Skilled Nursing Facilities Participating in the Pharmacy Partnership for Long-Term Care Program — United States, December 2020–January 2021. MMWR Morb Mortal Wkly Rep 2021; 70. Available at: https://www.cdc.gov/mmwr/volumes/70/wr/mm7005e2.htm. Accessed 27 April 2021.

5. Conlen M, Mervosh S, Ivory D. Nursing Homes, Once Hotspots, Far Outpace U.S. in Covid Declines. N. Y. Times. 2021; Available at: https://www.nytimes.com/interactive/2021/02/25/us/nursing-home-covid-vaccine.html. Accessed 27 April 2021.

6. Britton A. Effectiveness of the Pfizer-BioNTech COVID-19 Vaccine Among Residents of Two Skilled Nursing Facilities Experiencing COVID-19 Outbreaks — Connecticut, December 2020–February 2021. MMWR Morb Mortal Wkly Rep 2021; 70. Available at: https://www.cdc.gov/mmwr/volumes/70/wr/mm7011e3.htm. Accessed 29 April 2021.

7. Kuehn BM. Israel’s Real-life Evidence That Vaccine Can Prevent Severe COVID-19. JAMA 2021; 325:1603–1603.

8. Walsh EE, Frenck RW, Falsey AR, et al. Safety and Immunogenicity of Two RNA-Based Covid-19 Vaccine Candidates. N Engl J Med 2020; 383:2439–2450.

9. Anderson EJ, Rouphael NG, Widge AT, et al. Safety and Immunogenicity of SARS-CoV-2 mRNA-1273 Vaccine in Older Adults. N Engl J Med 2020; 383:2427–2438.

10. Prendecki M, Clarke C, Brown J, et al. Effect of previous SARS-CoV-2 infection on humoral and T-cell responses to single-dose BNT162b2 vaccine. Lancet Lond Engl 2021;

11. Müller L, Andrée M, Moskorz W, et al. Age-dependent immune response to the Biontech/Pfizer BNT162b2 COVID-19 vaccination. Clin Infect Dis Off Publ Infect Dis Soc Am 2021;

12. Dagan N, Barda N, Kepten E, et al. BNT162b2 mRNA Covid-19 Vaccine in a Nationwide Mass Vaccination Setting. N Engl J Med 2021;

13. Yelin I, Katz R, Herzel E, et al. Associations of the BNT162b2 COVID-19 vaccine effectiveness with patient age and comorbidities. medRxiv 2021; :2021.03.16.21253686.

14. Moustsen-Helms IR, Emborg H-D, Nielsen J, et al. Vaccine effectiveness after 1st and 2nd dose of the BNT162b2 mRNA Covid-19 Vaccine in long-term care facility residents and healthcare workers – a Danish cohort study. medRxiv 2021; :2021.03.08.21252200.

15. Chen RE, Zhang X, Case JB, et al. Resistance of SARS-CoV-2 variants to neutralization by monoclonal and serum-derived polyclonal antibodies. Nat Med 2021; :1–10.

16. Wang P, Nair MS, Liu L, et al. Antibody Resistance of SARS-CoV-2 Variants B.1.351 and B.1.1.7. Nature 2021; :1–9.

17. Zhou D, Dejnirattisai W, Supasa P, et al. Evidence of escape of SARS-CoV-2 variant B.1.351 from natural and vaccine-induced sera. Cell 2021; 184:2348-2361.e6.

18. Hacisuleyman E, Hale C, Saito Y, et al. Vaccine Breakthrough Infections with SARS-CoV-2 Variants. N Engl J Med 2021; 0:null.

19. Teran RA. Postvaccination SARS-CoV-2 Infections Among Skilled Nursing Facility Residents and Staff Members — Chicago, Illinois, December 2020–March 2021. MMWR Morb Mortal Wkly Rep 2021; 70. Available at: https://www.cdc.gov/mmwr/volumes/70/wr/mm7017e1.htm. Accessed 29 April 2021.

20. Manisty C, Otter AD, Treibel TA, et al. Antibody response to first BNT162b2 dose in previously SARS-CoV-2-infected individuals. Lancet Lond Engl 2021; 397:1057–1058.

21. Krammer F, Srivastava K, Alshammary H, et al. Antibody Responses in Seropositive Persons after a Single Dose of SARS-CoV-2 mRNA Vaccine. N Engl J Med 2021; 384:1372–1374.

22. Stamatatos L, Czartoski J, Wan Y-H, et al. mRNA vaccination boosts cross-variant neutralizing antibodies elicited by SARS-CoV-2 infection. Science 2021;

23. Samanovic MI, Cornelius AR, Wilson JP, et al. Poor antigen-specific responses to the second BNT162b2 mRNA vaccine dose in SARS-CoV-2-experienced individuals. MedRxiv Prepr Serv Health Sci 2021;

24. Lustig Y, Nemet I, Kliker L, et al. Neutralizing Response against Variants after SARS-CoV-2 Infection and One Dose of BNT162b2. N Engl J Med 2021; 0:null.

25. Ebinger JE, Fert-Bober J, Printsev I, et al. Antibody responses to the BNT162b2 mRNA vaccine in individuals previously infected with SARS-CoV-2. Nat Med 2021;

26. Outlining the Prior Infection with SARS-CoV-2 study (PICOV) -preliminary findings on symptoms in nursing home residents and staff. 2021. Available at: https://www.researchsquare.com. Accessed 9 April 2021.

27. Mariën J, Ceulemans A, Michiels J, et al. Evaluating SARS-CoV-2 spike and nucleocapsid proteins as targets for antibody detection in severe and mild COVID-19 cases using a Luminex bead-based assay. J Virol Methods 2021; 288:114025.

28. Earle KA, Ambrosino DM, Fiore-Gartland A, et al. Evidence for antibody as a protective correlate for COVID-19 vaccines. medRxiv 2021; :2021.03.17.20200246.

29. Vogel AB, Kanevsky I, Che Y, et al. BNT162b vaccines protect rhesus macaques from SARS-CoV-2. Nature 2021; 592:283–289.

30. Corbett KS, Flynn B, Foulds KE, et al. Evaluation of the mRNA-1273 Vaccine against SARS-CoV-2 in Nonhuman Primates. N Engl J Med 2020; 383:1544–1555.

31. McMahan K, Yu J, Mercado NB, et al. Correlates of protection against SARS-CoV-2 in rhesus macaques. Nature 2021; 590:630–634.

32. Parry HM, Tut G, Faustini S, et al. BNT162b2 Vaccination in People Over 80 Years of Age Induces Strong Humoral Immune Responses with Cross Neutralisation of P.1 Brazilian Variant. Rochester, NY: Social Science Research Network, 2021. Available at: https://papers.ssrn.com/abstract=3816840. Accessed 8 April 2021.

33. Long Q-X, Tang X-J, Shi Q-L, et al. Clinical and immunological assessment of asymptomatic SARS-CoV-2 infections. Nat Med 2020; 26:1200–1204.

34. Keehner J, Horton LE, Pfeffer MA, et al. SARS-CoV-2 Infection after Vaccination in Health Care Workers in California. N Engl J Med 2021; 0:null.

35. Kustin T, Harel N, Finkel U, et al. Evidence for increased breakthrough rates of SARS-CoV-2 variants of concern in BNT162b2 mRNA vaccinated individuals. medRxiv 2021; :2021.04.06.21254882.

36. COVID-19 Breakthrough Case Investigations and Reporting | CDC. 2021. Available at: https://www.cdc.gov/vaccines/covid-19/health-departments/breakthrough-cases.html. Accessed 5 May 2021.

37. Tsang JS, Dobaño C, VanDamme P, et al. Improving Vaccine-Induced Immunity: Can Baseline Predict Outcome? Trends Immunol 2020; 41:457–465.

38. Kotliarov Y, Sparks R, Martins AJ, et al. Broad immune activation underlies shared set point signatures for vaccine responsiveness in healthy individuals and disease activity in patients with lupus. Nat Med 2020; 26:618–629.

