## Supplementary material for "Poor antibody response to BioNTech/Pfizer COVID-19 vaccination in SARS-CoV-2 naïve residents of nursing homes"

**Supplemental Table S1. Outcome measurements according to study group and time point (day 0, 21, 28, and 49).** Statistical comparisons were made between the different participant groups using the Kruskall-Wallis test by ranks.

| | naive staff (N=19) | naive resident (N=20) | infected staff (N=21) | infected resident (N=20) | Total (N=80) | p value |
|---|---|---|---|---|---|---|
| **Binding RBD-IgG (AU/mL)** | | | | | | |
| **Day 0** | | | | | | 0.002 |
| Mean (SD) | 3.5 (4.7) | 1.1 (1.7) | 122.2 (110.4) | 284.1 (499.3) | 104.2 (276.5) | |
| Range | 0.1 - 19.0 | 0.1 - 5.0 | 3.0 - 528.0 | 13.0 - 2275.0 | 0.1 - 2275.0 | |
| **Day 21** | | | | | | <0.001 |
| Mean (SD) | 296.2 (169.7) | 42.3 (92.7) | 6910.4 (3127.7) | 6178.7 (4139.6) | 3439.6 (4123.9) | |
| Range | 34.0 - 659.0 | 0.1 - 425.0 | 245.0 - 12417.0 | 583.0 - 12950.0 | 0.1 - 12950.0 | |
| **Day 28** | | | | | | <0.001 |
| Mean (SD) | 2105.2 (1122.4) | 355.0 (447.0) | 8083.9 (2748.3) | 8867.0 (4314.4) | 4927.5 (4528.9) | |
| Range | 503.0 - 3619.0 | 0.1 - 1590.0 | 1683.0 - 14033.0 | 1141.0 - 14900.0 | 0.1 - 14900.0 | |
| **Day 49** | | | | | | <0.001 |
| Mean (SD) | 2058.3 (1363.7) | 1027.5 (946.4) | 8065.8 (3089.4) | 13102.0 (6781.1) | 6050.4 (6104.4) | |
| Range | 487.0 - 4472.0 | 53.0 - 3084.0 | 1552.0 - 13415.0 | 2029.0 - 26849.0 | 53.0 - 26849.0 | |
| **Binding S1-IgG (AU/mL)** | | | | | | |
| **Day 0** | | | | | | <0.001 |
| Mean (SD) | 1.6 (2.3) | 1.7 (3.5) | 71.6 (56.6) | 122.5 (105.5) | 50.2 (78.1) | |
| Range | 0.1 - 9.0 | 0.1 - 15.0 | 0.1 - 255.0 | 5.0 - 345.0 | 0.1 - 345.0 | |
| **Day 21** | | | | | | <0.001 |
| Mean (SD) | 172.5 (114.5) | 21.6 (34.9) | 6296.5 (3379.3) | 5557.5 (4198.8) | 3088.6 (3979.9) | |
| Range | 22.0 - 371.0 | 0.1 - 140.0 | 120.0 - 13058.0 | 527.0 - 14447.0 | 0.1 - 14447.0 | |
| **Day 28** | | | | | | <0.001 |
| Mean (SD) | 1037.3 (640.4) | 193.7 (263.0) | 7250.7 (3260.1) | 8226.1 (4977.4) | 4254.6 (4669.4) | |
| Range | 294.0 - 2145.0 | 0.1 - 1038.0 | 665.0 - 13759.0 | 829.0 - 17270.0 | 0.1 - 17270.0 | |
| **Day 49** | | | | | | <0.001 |
| Mean (SD) | 1299.3 (1012.0) | 509.1 (525.7) | 6134.2 (2927.9) | 10649.2 (5663.1) | 4633.2 (5137.5) | |
| Range | 246.0 - 4195.0 | 31.0 - 2211.0 | 925.0 - 12703.0 | 1326.0 - 19822.0 | 31.0 - 19822.0 | |

*Table continued on the next page*

### Supplemental Table S1 (continued)

| | naive staff (N=19) | naive resident (N=20) | infected staff (N=21) | infected resident (N=20) | Total (N=80) | p value |
|---|---|---|---|---|---|---|
| **Binding S2-IgG (AU/mL)** | | | | | | |
| **Day 0** | | | | | | <0.001 |
| Mean (SD) | 9.1 (6.2) | 4.0 (4.4) | 83.4 (54.2) | 113.6 (93.1) | 53.4 (71.3) | |
| Range | 0.1 - 21.0 | 0.1 - 13.0 | 17.0 - 220.0 | 11.0 - 414.0 | 0.1 - 414.0 | |
| **Day 21** | | | | | | <0.001 |
| Mean (SD) | 41.8 (29.3) | 20.5 (53.9) | 1578.8 (1196.6) | 1184.1 (1516.6) | 725.5 (1183.9) | |
| Range | 4.0 - 92.0 | 0.1 - 248.0 | 54.0 - 3657.0 | 155.0 - 6114.0 | 0.1 - 6114.0 | |
| **Day 28** | | | | | | <0.001 |
| Mean (SD) | 131.4 (98.2) | 25.6 (42.7) | 1418.1 (968.6) | 1476.7 (1702.6) | 779.1 (1189.0) | |
| Range | 6.0 - 357.0 | 0.1 - 200.0 | 98.0 - 3892.0 | 150.0 - 5547.0 | 0.1 - 5547.0 | |
| **Day 49** | | | | | | <0.001 |
| Mean (SD) | 77.1 (80.4) | 42.4 (47.0) | 718.0 (499.9) | 717.2 (748.1) | 392.6 (552.4) | |
| Range | 5.0 - 329.0 | 4.0 - 218.0 | 125.0 - 2307.0 | 176.0 - 3434.0 | 4.0 - 3434.0 | |
| **RBD-IgG avidity $K_{off}$ (1/s)** | | | | | | |
| **Day 0** | | | | | | |
| Not measured | 19 | 20 | 8 | 3 | 50 | |
| Mean (SD) | . | . | 3.2e-03 (2.7e-03) | 2.3e-04 (2.5e-03) | 2.7e-03 (2.6e-03) | |
| Range | . | . | 4.9e-04 - 9.6e-03 | 1.3e-04 - 7.9e-03 | 1.3e-04 - 9.6e-03 | |
| **Day 21** | | | | | | <0.001 |
| Not measured | 3 | 16 | . | . | 18 | |
| Mean (SD) | 8.2e-04 (6.2e-04) | 2.7e-03 (2.4e-03) | 7.5e-05 (1.2e-04) | 6.3e-05 (5.7e-05) | 4.5e-04 (9.3e-04) | |
| Range | 2.8e-04 - 2.5e-03 | 5.4e-04 - 4.9e-03 | 5.5e-06 - 4.4e-04 | 1.9e-07 - 2.1e-04 | 1.9e-07 - 4.9e-03 | |
| **Day 28** | | | | | | <0.001 |
| Not measured | 1 | 6 | . | . | 7 | |
| Mean (SD) | 1.5e-04 (7.0e-05) | 1.3e-03 (2.3e-03) | 6.1e-05 (1.2e-04) | 4.8e-05 (3.7e-05) | 3.2e-04 (1.1e-03) | |
| Range | 7.5e-05 - 3.5e-04 | 2.2e-04 - 8.9e-03 | 1.5e-06 - 5.4e-04 | 1.0e-07 - 1.6e-04 | 1.0e-07 - 8.9e-03 | |
| **Day 49** | | | | | | <0.001 |
| Not measured | . | 5 | . | . | 6 | |
| Mean (SD) | 2.3e-04 (1.7e-04) | 4.8e-04 (4.2e-04) | 6.0e-05 (8.8e-05) | 3.4e-05 (3.3e-05) | 1.8e-04 (2.7e-04) | |
| Range | 8.6e-05 - 6.9e-04 | 1.1e-04 - 1.8e-03 | 1.0e-07 - 4.0e-04 | 1.0e-07 - 1.3e-04 | 1.0e-07 - 1.8e-03 | |

*Table continued on the next page*

**Supplemental Table S1 (continued)**

| | naive staff (N=19) | naive resident (N=20) | infected staff (N=21) | infected resident (N=20) | Total (N=80) | p value |
|---|---|---|---|---|---|---|
| **50% neutralization (wild type)** | | | | | | |
| **Day 0** | | | | | | 0.17 |
| Mean (SD) | 25.0 (0.0) | 25.0 (0.0) | 142.4 (375.7) | 104.5 (109.6) | 75.7 (203.2) | |
| Range | 25.0 - 25.0 | 25.0 - 25.0 | 25.0 - 1759.7 | 25.0 - 459.5 | 25.0 - 1759.7 | |
| **Day 21** | | | | | | <0.001 |
| Mean (SD) | 29.8 (11.7) | 25.0 (0.0) | 4218.5 (2592.0) | 4696.8 (4480.8) | 2294.9 (3393.0) | |
| Range | 25.0 - 65.3 | 25.0 - 25.0 | 25.0 - 10473.0 | 400.0 - 16726.2 | 25.0 - 16726.2 | |
| **Day 28** | | | | | | <0.001 |
| Mean (SD) | 311.7 (221.4) | 47.7 (39.7) | 5109.8 (2927.3) | 6036.5 (4902.4) | 2936.4 (3926.7) | |
| Range | 25.0 - 635.0 | 25.0 - 132.9 | 134.6 - 11143.0 | 348.2 - 22286.1 | 25.0 - 22286.1 | |
| **Day 49** | | | | | | <0.001 |
| Mean (SD) | 222.4 (198.8) | 70.7 (61.2) | 5126.2 (3837.2) | 4616.5 (4436.7) | 2544.3 (3748.4) | |
| Range | 25.0 - 700.0 | 25.0 - 237.8 | 87.4 - 14452.0 | 400.0 - 19590.8 | 25.0 - 19590.8 | |
| **50% neutralization (B.1.351)** | | | | | | |
| **Day 0** | | | | | | 0.72 |
| Not measured | 17 | 11 | 9 | 3 | 40 | |
| Mean (SD) | 25.0 (0.0) | 25.0 (0.0) | 25.0 (0.0) | 657.6 (2537.1) | 293.9 (1655.6) | |
| Range | 25.0 - 25.0 | 25.0 - 25.0 | 25.0 - 25.0 | 25.0 - 10500.3 | 25.0 - 10500.3 | |
| **Day 21** | | | | | | 0.006 |
| Not measured | 11 | 11 | 2 | . | 24 | |
| Mean (SD) | 25.0 (0.0) | 25.0 (0.0) | 1046.9 (1013.1) | 891.8 (1059.7) | 687.7 (961.0) | |
| Range | 25.0 - 25.0 | 25.0 - 25.0 | 174.1 - 4271.5 | 25.0 - 3200.0 | 25.0 - 4271.5 | |
| **Day 28** | | | | | | <0.001 |
| Not measured | . | 4 | . | . | 4 | |
| Mean (SD) | 42.8 (20.1) | 25.0 (0.0) | 1164.5 (1214.6) | 1318.8 (1310.9) | 684.8 (1094.4) | |
| Range | 25.0 - 81.8 | 25.0 - 25.0 | 25.0 - 5011.1 | 25.0 - 3200.0 | 25.0 - 5011.1 | |
| **Day 49** | | | | | | <0.001 |
| Mean (SD) | 29.2 (12.5) | 25.0 (0.0) | 637.2 (483.0) | 1282.3 (1536.2) | 491.1 (932.9) | |
| Range | 25.0 - 66.4 | 25.0 - 25.0 | 25.0 - 1806.5 | 25.0 - 5207.8 | 25.0 - 5207.8 | |

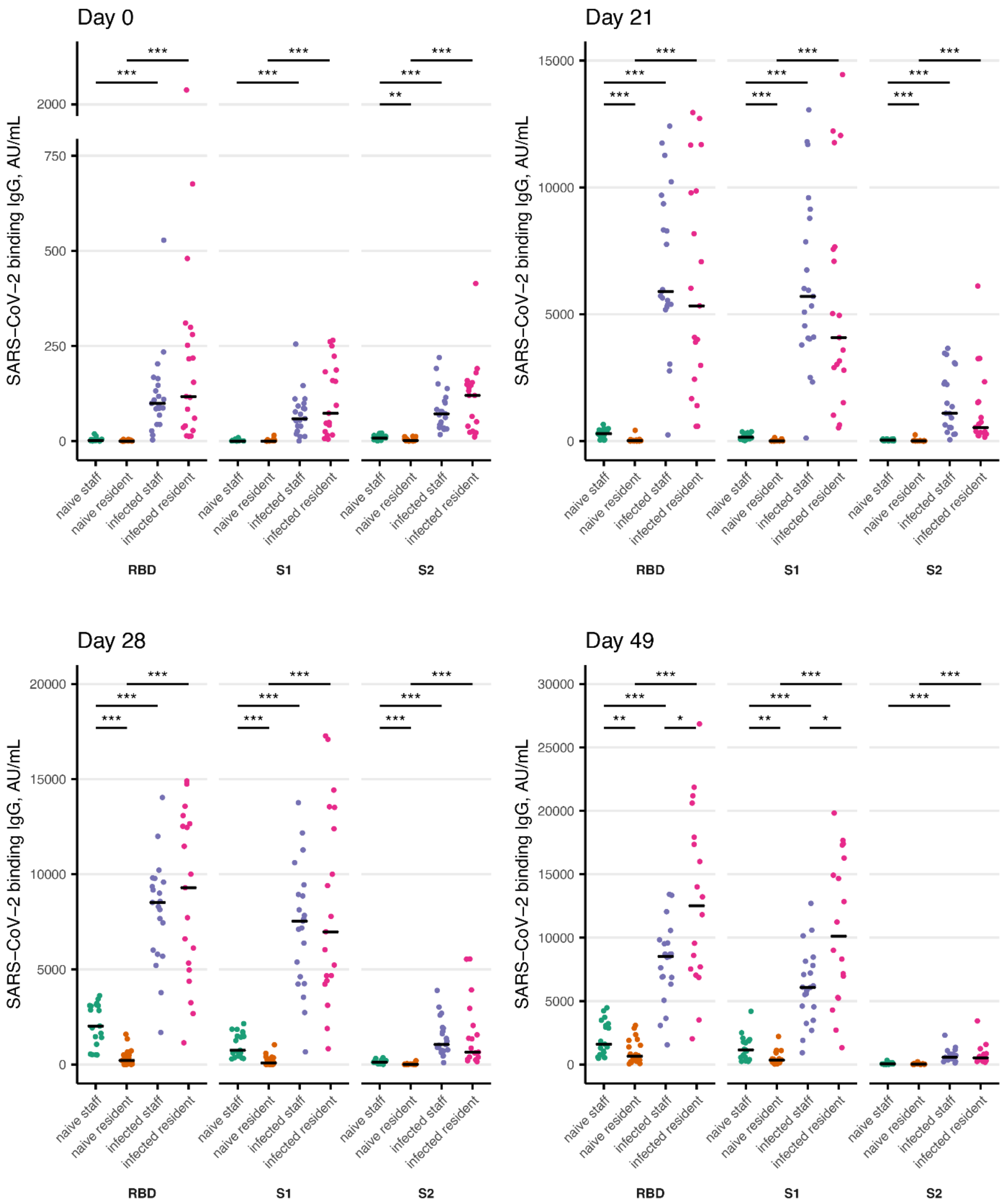


**Supplemental Figure S1. Multiplex immunoassay results at the different sampling times.**

The spike binding IgG antibodies (RBD, S1, and S2) are presented for each sampling day. Statistical comparisons were made between the different participant groups using the Kruskall-Wallis test by ranks, and the Mann-Whitney U post-hoc test. Statistical significance is denoted as: * p<0.05; ** p<0.01; and *** p<0..001

**Supplemental methods. Cutoff determination for anti-RBD IgG, anti-S1 IgG and anti-S2 IgG**

For establishing reliable cutoff values for RBD, S1, and S2, sera of 184 PCR-confirmed COVID-19 patients (collected >14 days post onset of symptoms, comprising mild (n=111) and severe (n=73) clinical outcomes) were tested in the Multiplex SARS-CoV-2 Immunoassay (serum dilution 1/100) and the results compared to a control panel of 259 sera, comprising PCR-negative (n=93) and pre-pandemic (n=166) sera. Assay performance at each individual cutoff was evaluated using ROC analyses and a specificity-optimized cutoff (specificity of at least 98.5%) was determined for each antigen. For S1, values are largely overlapping between groups. To be able to set a reasonable cutoff for S1, ROC analysis for S1 was performed omitting values >20 AU/ml in the control group (presumably positive due to cross-reactivity with endemic human coronaviruses) and <15 AU/ml in the COVID-19 group (presumably negative due to unresponsiveness or waning). The ROC-analyses generated cutoff concentrations of 21.0 AU/ml, 19.5 AU/ml and 19.5 AU/ml for RBD, S1 and S2, respectively. These cutoffs resulted in a specificity of 100% (95%CI 98.54-100), 100% (95%CI 98.48-100) and 100% (95%CI 98.54-100) at a sensitivity of 100% (95% CI 97.95-100), 86.82% (95%CI 79.91-91.61) and 94.02% (95%CI 89.61-96.63) for RBD, S1 and S2, respectively.

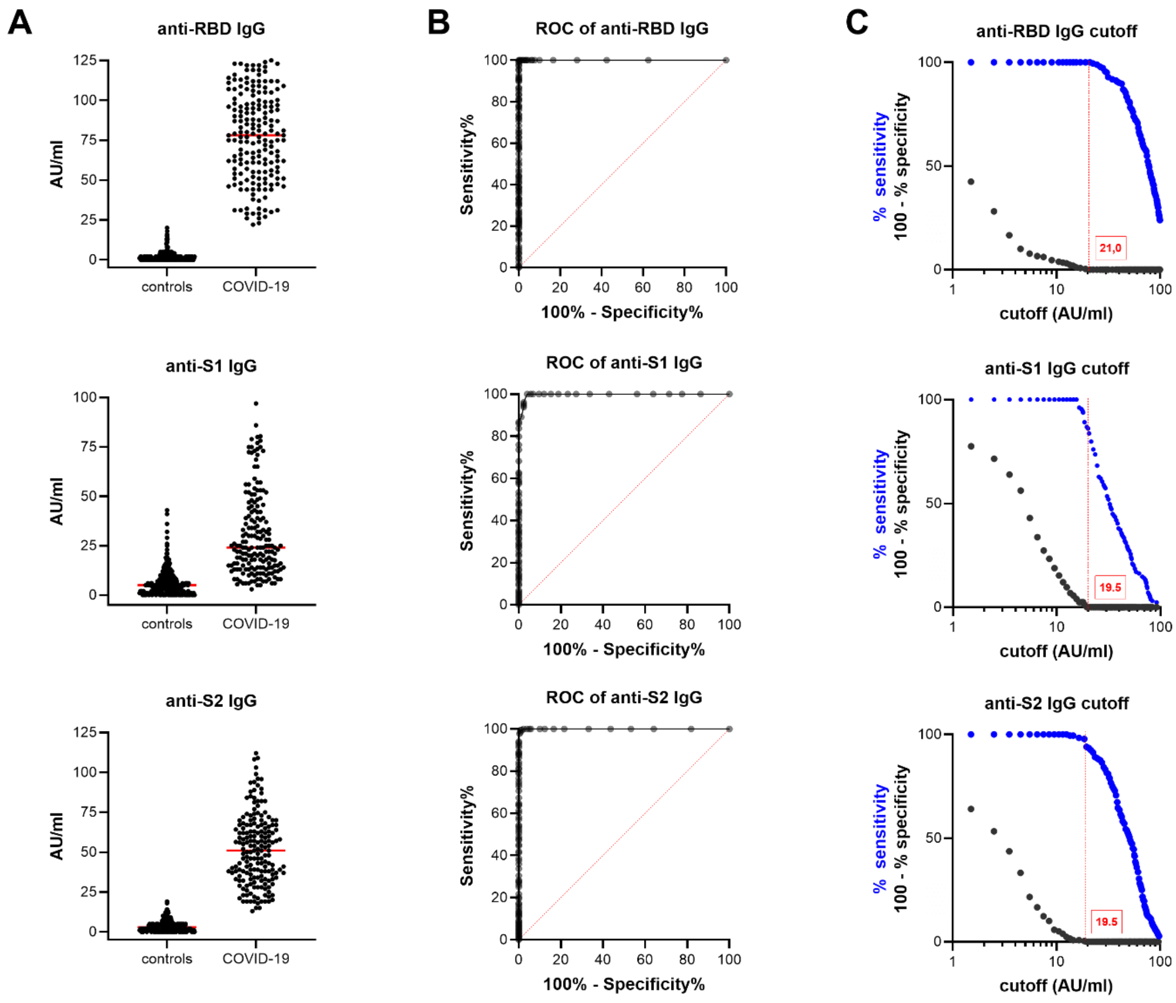


***Cutoff determination for anti-RBD IgG, anti-S1 IgG and anti-S2 IgG.*** *A, Concentrations of IgG were compared between control sera (n=259, PCR-negative and pre-pandemic) and COVID-19 sera (n=184, collected >14 days post onset of symptoms, mild and severe disease outcome). Median concentration is shown in red. B, ROC-analyses for RBD and S2 comprise all sera tested in (A)(n=443). For S1, ROC analysis was performed on 378 samples, excluding presumably positive and negative sera. C, The ROC data were used to determine a specificity-optimized cutoff of at*

*least 98.5% specificity (red dashed line). Abbreviations: AU, arbitrary unit; IgG, immunoglobulin G; RBD, receptor binding domain; S1, spike protein subunit 1; S2, spike protein subunit 2; ROC, receiver operator characteristic.*
